# The African American Dementia and Aging Project (AADAPt): An Oregon-based Longitudinal Study

**DOI:** 10.1101/2024.05.06.24306831

**Authors:** Allison Lindauer, Raina Croff, Kevin Duff, Nora Mattek, Patrice Fuller, Aimee Pierce, Kalisha Bonds, Jeffrey Kaye

## Abstract

**Objectives:** The vast majority of studies on aging, cognition, and dementia focus on non-Hispanic white subjects. This paper adds to the extant literature by providing insight into the African American aging experience. Here we describe the study design and baseline characteristics of the African American Dementia and Aging Project (AADAPt) study, which is exploring aging and cognition in African American older adults in Oregon.

**Methods:** African American older adults (n=177) participated in AADAPt, a longitudinal study that collected data on cognitive, physical, and social functioning in annual visits since 2000.

**Results:** AADAPt participants had risk factors for developing dementia in future, such as hypertension and hyperlipidemia, but also reported protective factors such as high social engagement.

**Conclusions:** The AADAPt project offers new insights into aging in older African Americans that includes data on cognition, social engagement, and physical health, which are crucial for understanding the experience of under-represented groups and making future studies more inclusive. These findings reflect a window of time for a geographically-focused cohort, and the lessons learned from this study likely have broader implications for shaping the health of these older African American adults. Keywords: African American, Dementia, Observational Study

## Introduction

Although one in nine older Americans has dementia due to Alzheimer’s disease and related dementias (ADRD) (Alzheimer’s Association, 2024), African Americans bear a heavier burden due to the significantly higher prevalence of ADRD in this population (Mehta & Yeo, 2017; Plassman et al., 2007). However, our understanding of cognitive and functional declines in the African American population remains limited. The African American Dementia and Aging Project (AADAPt), housed within the Oregon Alzheimer’s Disease Research Center (OADRC), aims to explore and define the physical, cognitive, and social factors that contribute to health and cognitive decline in this historically underrepresented group. Our long-term goal is to use such findings to develop predictive models of aging and cognitive and functional decline in this sample, as well as inform the methods and processes that are necessary for engaging these individuals in observational and interventional research. This paper describes the study design and baseline data, setting the stage for future analyses.

It has been posited that the risk of ADRD in African Americans is higher due to physical and medical conditions such as hypertension (Mayeda et al., 2014; Schneider et al., 2015), and to historical and ongoing systemic inequities such as educational quality. For example, cross-sectional studies indicate that lower educational attainment in African Americans is associated with dementia, (Callahan et al., 1996; Hall et al., 2000), but several longitudinal studies found the association between education, race, and cognitive decline to be more nuanced (Carvalho et al., 2015; Sisco et al., 2015). A few studies have examined how other factors, such as social engagement (Barnes et al., 2004; Hamlin et al., 2022) and physical function (Studenski et al., 2011; Wolinsky et al., 2014) affect the African American aging process and cognitive well-being over time. To further illuminate the impact of ADRD on African Americans, the overarching aims of AADAPt are to: 1) identify predictors of successful aging in this Oregon-based African American community, and 2) elucidate factors that contribute to cognitive decline in the group.

To our knowledge, this is the only longitudinal study of African Americans and aging in Oregon. The Oregon perspective offers a unique opportunity to understand how time, place, and education affect aging and cognitive function over time. The African American community is relatively new to this part of the United States. Oregon’s first constitution banned non-white citizens from the state but this was reversed by the time transcontinental train systems extended to the Pacific Northwest in the early 1900s (Brooks, 2006; Oregon State Constitution, 1857). The number of African Americans in Oregon was small until World War II (WWII), when the high demand for shipyard workers brought thousands to the region (Demarco, 1990; McLagen, 1980). Many lived in a company town which did not segregate neighborhoods or schools (Maben, 2000).

While Oregon skirted the oppressive Jim Crow laws of the mid-20^th^ century, the African American community still faced considerable discrimination and racism in the Pacific Northwest (McLagen, 1980). The Ku Klux Klan had an early influential presence in Oregon, with up to 25,000 members in 1922 (Chalmers, 1965; Richard, 1983). Later, at the conclusion of WWII, many African American citizens faced challenges finding housing due to strict “redlining” laws, in which options were limited to circumscribed areas in Portland, Oregon’s largest city (Holmes, 1948). In response to the racist mortgage lending practices, the vast majority of Portland’s African American community settled in the cluster of conjoined neighborhoods, known as the Albina District (Bates 2019; Gibson 2007).

Over the years, the African American community’s response to discrimination in the Pacific Northwest has been solidarity. Social networks were built and strengthened through memberships in clubs, churches, workers’ unions, and other social venues, reinforcing the strength of the community while simultaneously defining it on its own terms rather than on those of the dominant culture (Bates, 2019; Millner, 1995). Neighbors grew up with intergenerational knowledge of one another, cultivating long-term trusted bonds (Croff et al., 2021). Close proximity of resources, including social support, is critical for aging in place (Levasseur et al., 2015), and living within the Albina District’s concentrated Black community meant that older adults and their families could draw from a deep and wide communal pool of close-proximity familial and neighborly support, consistent with Black family dementia caregiving ecosystems and values of minimal formal care usage (Bonds Johnson et al., 2022; Brewster et al., 2020). A strong sense of social support and opportunities for neighborhood-based social engagement help mitigate family burden and cognitive decline, and counter depression and anxiety (Brewster et al., 2020; Clay et al., 2008).

More recently, gentrification-related community displacement is fracturing close-proximity social supports, decreasing social networks, and increasing stress (Croff et al., 2021; Smith et al., 2018; Versey, 2018). These changes can be detrimental for older Black families who remain in gentrifying neighborhoods and potentially for those who relocate. Thus, it remains to be seen if gentrification in Portland’s historically African American neighborhoods and the related dispersal of residents will influence the cognitive health and rates of ADRD of AADAPt participants. Considering this context, the AADAPt study may provide valuable insight on the potential effects of temporal demographic and social trends on aging and dementia.

## Methods

### Design and setting

This longitudinal, observational study assesses the physical, cognitive, and social health of older African American participants annually. Recruitment is continuous, with the first participants enrolled in 2000. Here we describe the sample and findings up to 2019. The protocol is approved by the OHSU Institutional Review Board (IRB #00001480) and all participants provided written informed consent prior to engaging in study procedures. Data is available from our local repository (#6845), and is submitted to the National Alzheimer’s Coordinating Center Repository, which adds regional diversity to the national sample (https://naccdata.org/requesting-data/nacc-data).

### Participants

Participants (n=177) were recruited from the Portland, Oregon metropolitan area. To facilitate recruitment, the AADAPt research team collaborates with local community organizations, such as PreSERVE Coalition (https://www.preserve-coalition.org ), a group of community members and academicians that promote brain health in the African American community. Churches, sororities, and health care systems also promote recruitment opportunities by providing space for information sessions, study presentations, and networking opportunities.

To be included in the study, participants need to self-identify as Black/African American, age 55 years or more. They cannot have a dementia diagnosis at study entry, and they need to able to participate in cognitive and functional testing. They must have a study partner to provide collateral historical data. Participants are encouraged to continue with the study for as long as they are able.

To encourage participation and reduce barriers to enrollment, transportation to research visits is provided free of charge. Participants are remunerated $30 per visit for their time and effort. Participants are thanked and updated on study findings at an annual brunch hosted by the research team in the community’s neighborhood.

### Procedures and measures

Enrolled participants attend clinic-based visits annually. The health history of participants, including family history of dementia, is documented. They are offered the option of assenting to brain donation after death for genetic and other analyses.

During the annual visits, a broad range of physical, cognitive, and social data is collected on each participant (see Table 1). These AADAPt participants had the option of receiving a brain MRI for volumetric analyses (Mueller et al., 1998), and blood samples for apolipoprotein E evaluation (Farrer et al., 1997).

**Table 1.**
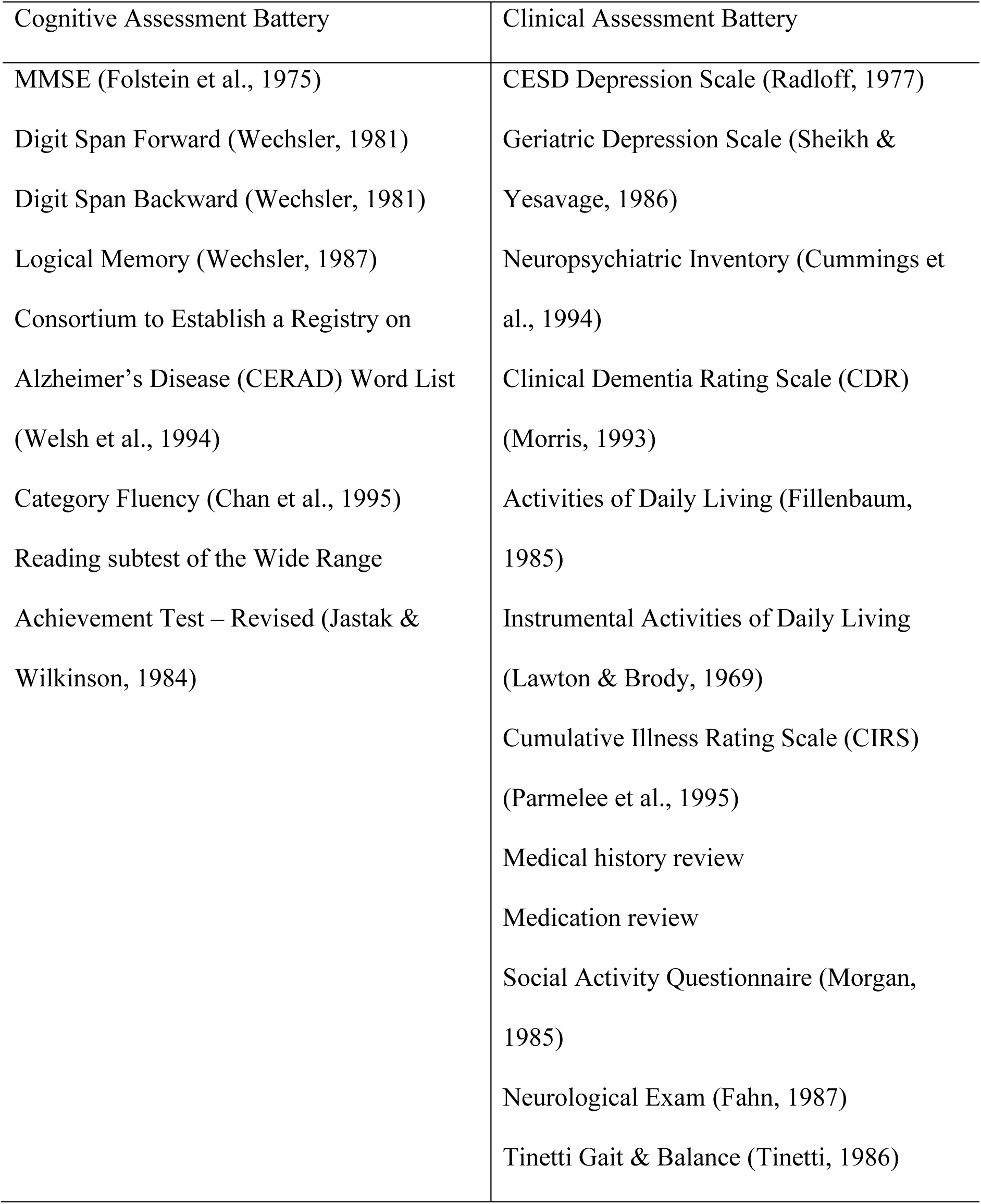
AADAPt Baseline Assessment Batteries.

### Statistical analyses

Baseline descriptive findings of health-related risk factors (e.g., hypertension, diabetes), cognition, and social functioning are presented for the overall cohort. Additionally, comparisons were made between those classified as cognitively and functioning intact at baseline (i.e., Global Clinical Dementia Rating (CDR)=0, (Morris, 1993)) and those with at least questionable cognition and/or daily functioning (i.e., CDR>0) with a series of three MANCOVAs that controlled for group differences on age and education: 1) Daily functioning (i.e., basic and instrumental activities of daily living (Fillenbaum, 1985; Lawton & Brody, 1969)), 2) cognition (i.e., demographically-corrected z-scores on WRAT-R Reading, MMSE, Digit Span Forward and Backward, Category Fluency, CERAD Word List Acquisition and Delay Recall, Logical Memory I and II (Shirk et al., 2011; Weintraub et al., 2018; Weintraub et al., 2009)), and 3) physical functioning (i.e., CIRS (Parmelee et al., 1995), Tinetti Gait and Balance(Tinetti, 1986)). For these MANCOVAs, follow-up ANCOVAs examined which measures within the domain were significantly different between the two groups. Finally, two ANCOVAs, which also controlled for age and education, compared the two groups on 1) depression (i.e., GDS/CESD(Radloff, 1977; Sheikh & Yesavage, 1986)) and 2) social activity (i.e., a composite measure of social activities (Morgan, 1985)). Since data on this cohort was collected over a long period of time, this was considered as another possible covariate. However, there were not group differences (i.e., Global CDR=0 vs. >0) on time of data collection (p=0.08). Comparisons were considered statistically significantly different if p<0.05. All analyses were performed using SPSS 29.0 software.

## Results

### Demographics

The sample included 177 participants who met the inclusion criteria and completed a baseline visit (Table 2). Most were cognitively intact at enrollment (76%, Clinical Dementia Rating Scale=0, Morris 1993), and, based on responses from a collateral source, the participants were quite intact for basic and instrumental activities of daily living. Nearly half of the participants lived alone at the time of the baseline evaluation (45%), 37% lived with a spouse or partner, and 17% lived with a family member (e.g., children, grandchildren, sibling). Most participants were born in the southern United States (70%) and moved to the Pacific Northwest in their youth. Over a third of the cohort (37%) was born in Texas or Louisiana.

**Table 2.** Baseline demographic and clinical measures for the total sample by cognitive status.

|  | Total | CDR=0 | CDR>0 |
| --- | --- | --- | --- |
| N | 172* | 130 | 42 |
| Age (years) | 72.6 (7.4) | 71.3 (6.7) | 76.9 (7.8) |
| Sex (% female) | 73.3% | 74.6% | 69.0% |
| Race, African American | 100% | -- | -- |
| Education (years) | 13.7 (3.1) | 14.2 (2.9) | 12.1 (3.4) |
| Basic ADLs | 0.3 (0.7) | 0.1 (0.5) | 0.6 (1.0) |
| Instrumental ADLs | 0.3 (1.2) | 0.1 (0.6) | 0.9 (2.0) |
| MMSE (raw) | 27.7 (2.4) | 28.4 (1.5) | 25.8 (3.3) |
| WRAT Reading (z) | 0.1 (1.4) | 0.2 (1.4) | -0.3 (1.5) |
| Digit Span Forward (z) | -0.3 (0.5) | -0.3 (0.5) | -0.3 (0.5) |
| Digit Span Backward (z) | -0.7 (0.5) | -0.7 (0.6) | -0.8 (0.5) |
| Category Fluency (z) | 0.0 (0.9) | 0.1 (0.9) | -0.4 (0.9) |
| CERAD List Acquisition<br>(z) | 0.1 (1.1) | 0.3 (1.0) | -0.5 (1.2) |
| CERAD List Recall (z) | 0.0 (1.2) | 0.2 (1.2) | -0.7 (1.2) |
| Logical Memory I (z) | -0.1 (1.1) | 0.0 (1.0) | -0.6 (1.1) |
| Logical Memory II (z) | 0.1 (1.1) | 0.3 (1.0) | -0.5 (1.3) |
| CIRS | 21.4 (3.9) | 20.9 (3.6) | 22.9 (4.4) |
| Tinnetti Balance | 2.0 (3.0) | 1.4 (2.1) | 3.7 (4.7) |
| Tinnetti Gait | 0.6 (1.3) | 0.4 (0.9) | 1.4 (1.9) |
| CESD/GDS | 1.4 (1.8) | 1.1 (1.6) | 2.3 (2.2) |
| Social composite | 10.0 (3.0) | 10.3 (2.8) | 8.6 (3.2) |

\*5 participants did not have a CDR

### Cognition

The majority of the sample (76%) had a CDR global rating of 0.0, with 24% having a rating of 0.5 (and <1% had a CDR of 1.0, but had a MMSE score that allowed inclusion). For the entire sample, 7% rated their memory as “excellent,” 59% as “good,” 29% as “fair,” and 5% as “poor” or “very poor.” Global cognition, as measured by the MMSE (Folstein et al., 1975), was consistent with subjective data, mean=27.6 (SD 2.4), which falls above the conventional cutoff for dementia. Premorbid intellect was average (i.e., WRAT Reading), as was simple attention (i.e., Digit Span Forward), semantic fluency (i.e., Category Fluency), and verbal learning and memory (i.e., CERAD List Acquisition and Recall, Logical Memory I and II). Complex attention (i.e., Digit Span Backward) was low average.

### Physical Healt

Participants were optimistic about their physical health, with 71% rating their overall physical health as “good” or “excellent”, 26% viewed their health as “fair,” and 3% endorsed “poor” or “very poor”. Despite this optimistic view of their health, a sizable proportion reported current or past stroke or transient ischemic attack (26%), diabetes (33%), hyperlipidemia (54%) and hypertension (83%), and 50% reported ever smoking. Over half (58%) reported a family history of dementia. Even with these multiple vascular risk factors and dementia family history, baseline medical burden was comparable to another cohort of community dwelling older adults (Kaye et al., 2011) (CIRS: mean=21.4, SD=3.9 [55^th^ percentile of Kaye et al., 2011]). Their balance and gait were also comparable to community dwelling older adults (Tinetti Gait: median=0 [50^th^ percentile of Kaye et al., 2011], Tinetti Balance median=1.5, [∼50^th^ percentile of Kaye et al., 2011]).

### Mental Health

In this cohort, 53% reported past or current depression. However, on screening measures of depressive symptoms (Table 1), minimal such symptoms were endorsed. Similarly, in a subset of this cohort, their collaterals endorsed minimal neuropsychiatric symptoms in them on the Neuropsychiatric Inventory total score = 0.23 (0.83) (Cummings et al., 1994).

### Social Activity

Participants reported active social engagement. For example, the modal number of current close friends that participants reported was five. Most reported engaging in the following activities daily or weekly: attending church (76%), going out to eat (57%), and having visitors (54%). They followed the news, with 57% reading the newspaper or 92% listening to news on the radio/television on a daily basis. Over one-third of the cohort reported daily use of a computer. Social activity was also related to other outcomes. For example, using a composite of social activities greater social engagement was associated with better global cognition (MMSE: r=0.24, p=0.005), lower depression symptom scores (GDS/CESD: r=-0.21, p=0.01), and lower medical comorbidity (CIRS: r=-0.21, p=0.008).

### Physiologic assessments

In a subset of 88 participants who completed an MRI of the brain, the average hippocampal volume was 1.3 cc (SD=0.2), and total brain volume= 908 cc (SD=103), WMH 5.0 cc (SD=6.3) (Mueller et al., 1998). Apolipoprotein E was assessed in 121 of the AADAPt participants. Of these, 67.5% of the sample had no *ϵ*4 alleles, 30% of the sample had one *ϵ*4 allele, and 2.6% of the sample had two *ϵ*4 alleles (Farrer et al., 1997; Logue et al., 2023).

### Comparison between CDR Groups

When the individuals with intact CDR scores (CDR=0) were compared to those with questionable or more severe dementia (CDR>0), the intact participants were younger (p<0.001) and had more years of education (p<0.001). The groups did not differ by gender (p=0.48) (Table 2). As such, age and education were used as covariates in these group comparisons. In an initial MANCOVA on daily functioning, the two groups differed on collateral ratings of their abilities to perform activities of daily living, with the CDR=0 group performing significantly better than the CDR>0 group (F[2,145]=7.1, p=0.001). In follow-up ANCOVAs, this difference was present on both scales of basic (p<0.001) and instrumental (p=0.006) activities.

In a MANCOVA examining cognition, there were overall differences between these two groups (F[9,120]=4.0, p<0.001), with the CDR=0 group performing better than the CDR>0 group. In follow-up ANCOVAs, there were no group differences on premorbid intellect or simple and complex attention. Conversely, those rated as intact had significantly better global cognition (p<0.001), semantic fluency (p=0.047), list learning (p=0.004) and recall (p<0.001), and story learning (p=0.01) and recall (p<0.001) than those with impaired CDR scores. Such scores differences suggest that the impaired group presented with an amnestic cognitive profile that could indicate preclinical Alzheimer’s disease.

In a MANCOVA on physical functioning, there were significant group differences (F[3,144]=4.1, p=0.008). In follow-up ANCOVAs, those with CDR=0 had fewer medical comorbidities (CIRS: p=0.04), better balance (Tinnetti balance: p=0.01), and better gait (Tinnetti gait: p=0.003) compared to those with CDR>0. In an ANCOVA examining responses on self-reported depression screenings, those with CDR=0 endorsed significantly fewer such symptoms compared to those with CDR>0 (F[1,170]=28.5, p=0.003). Finally, in an ANCOVA using a composite of social activities, individuals rated as CDR>0 engaged in significantly fewer social activities than those rated as CDR=0 (F[1,150]=39.3, p=0.03).

## Discussion

Since the prevalence of ADRD is significantly higher in African Americans (Mehta & Yeo, 2017; Plassman et al., 2007) and the understanding of cognitive changes in this population remains limited, we present the baseline results of AADAPt, which explores and defines physical, cognitive, and social factors that contribute to health and cognitive decline in this Pacific Northwest urban community. These findings add to the existing literature on the complex interplay of variables that influence cognitive health and impairment in this historically underrepresented group in medical research. These baseline results suggest that many participants had optimistic perceptions of their health, yet objective and important risk factors that could predict future cognitive impairment and functional decline were present.

Nearly three-quarters of AADAPt participants self-rated their physical health as good or excellent. Despite this subjective report, the cohort also had high rates of hypertension (83%), hyperlipidemia (54%), and diabetes (33%), and over a quarter of the sample had a history of a stroke or TIA, which is much higher than the national prevalence of stroke in this group (Tsao et al., 2023). This constellation of risk factors suggests this group is vulnerable to developing vascular dementia and Alzheimer’s disease (Arfanakis et al., 2020).

Similarly, two-thirds of the cohort rated their memory as good to excellent, and nearly as many reported no worsening in their memory over the past year. Objective measures of cognition were consistent with this report, as global cognition, simple attention, semantic fluency, verbal learning and memory all fell into the average range. This suggests, even with higher risk of cognitive decline, the current enrollees remain resilient to this risk.

More than half of AADAPt participants endorsed past or current depression, however responses on a screening measure of depressive symptoms were indicative of minimal symptoms of depression. Participants reported active social engagement (e.g., regularly attending church, going out to eat, having visitors) and greater social engagement. They individuals also regularly read the newspaper, listen to news, and use a computer. These activities were correlated with higher cognition, lower depression scores and less overall medical burden, and may explain why these individuals report few difficulties with low mood symptoms.

Three-quarters of the AADAPt cohort was cognitively healthy at baseline (CDR 0) and one-quarter had MCI (mild cognitive impairment) or questionable dementia (CDR>0). Cognitively healthy participants had fewer medical comorbidities, better cognition, fewer symptoms of depression, and were engaged in more social activities than those with questionable dementia. Longitudinal changes in these two groups will be important to examine if participants convert to MCI or progress to ADRD.

Studies indicate that subjective memory complaint can predict future cognitive decline (Lee & Foster, 2023) and is associated with depression. Our findings of high stroke prevalence and depression in the AADAPt cohort suggest this group is particularly vulnerable to future cognitive impairment. However, despite these risk factors, the participants had high levels of perceived cognitive and physical health. Strong community cohesion, coupled with this optimism may have protective features that need to be explored.

This study offers insight into cognitive, physical and social well-being in this cohort of African Americans in Oregon. It is unique in that it is a longitudinal study that explores a range of risk factors for dementia with participants who, at baseline, are cognitively intact. The AADAPt participants are well-characterized with physical, cognitive and social measures collected annually. Our findings suggest that there is an important interplay between environment and perception, but how these relationships play out is unknown.

### Limitations

AADAPt participants were recruited from the metropolitan Portland area, which limits the geographic generalizability of these findings. Participants were also self-selected, which could create some bias in this sample. Our sample has limited representation from those with lower educational levels.

## Conclusions

African Americans have a greater prevalence of MCI and ADRD than the general population. Additional data is needed to better understand the risk factors associated with cognitive decline as well as the modifiable preventive factors that may be promoted before cognitive and functional deficits develop. This ongoing study will inform cognitive and functional changes in older, community-dwelling African Americans. As such, the AADAPt study provides a deeper understanding of factors that may influence cognitive health for older African Americans living in the Pacific Northwest.

## Data Availability

Data is available from the National Alzheimers Coordinating Center Repository.

https://naccdata.org/requesting-data/nacc-data

